# HPV-Stratified Causal Digital Twins for Personalized Survival Benefit Estimation in Head and Neck Cancer

**DOI:** 10.64898/2026.09.30.26363910

**Authors:** Keyur D. Shah, Chih-Wei Chang, Jun Zhou, Jie Ding, Anees Dhabaan, Hania A. Al-Hallaq, James Bates, William Stokes, David S. Yu, Xiaofeng Yang

## Abstract

**Purpose:** Digital-twin frameworks can potentially offer a pathway toward individualized treatment but remain largely untested in radiation oncology. We present a causal digital-twin model for head-and-neck (H&N) cancer that estimates patient-specific survival benefit from chemoradiotherapy (CRT) versus radiotherapy (RT) alone, stratified by HPV status.

**Methods:** Using the public RADCURE cohort (n = 3,346), we developed a causal digital-twin framework in which patients were divided into HPV-positive and HPV-negative groups and assigned to training (RT start year ≤ 2007) and test sets (RT start year > 2007). Bayesian-network structure learning identified each cohort’s Markov Blanket around the 2-year survival endpoint (SVy2), yielding the minimal causally relevant feature set. A causal-forest model (double machine learning, DML framework) estimated the individualized treatment effect *τ* = *P*(*Svy2* | *CRT,X*) − *P*(*Svy2* | *RT,X*), with 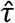 denoting the model-estimated effect. Overlap restrictions were applied to mitigate extrapolation. Predictive discrimination (AUC, C-index), average treatment effects (ATE), individualized effects (ITE), and decision-curve analyses were computed.

**Results:** For HPV-negative disease, internal evaluation demonstrated strong discriminative performance (test AUC = 0.72; C-index = 0.81, p < 0.01). The estimated test-set ATE favored CRT with a modest but positive improvement in SVy2 (mean 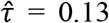). Patterns in 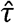 were clinically coherent, including increased benefit in node-positive and good-performance patients and reduced benefit in high-burden or poor-performance presentations. Decision-curve analysis showed that a 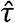-guided selective-CRT strategy provided higher net benefit than uniform treatment policies. HPV-positive disease exhibited substantially greater but less clinically structured treatment-effect heterogeneity, accompanied by reduced overlap and greater uncertainty.

**Conclusion:** This study demonstrates a proof-of-concept causal digital-twin framework for interpretable, patient-specific survival simulation in H&N cancer. The model captures clinically plausible treatment-effect heterogeneity and establishes a foundation for multi-cohort validation and future integration into individualized treatment decision support.

## Introduction

Head-and-neck (H&N) cancer management requires balancing potential survival benefit against treatment-related toxicity and patient burden. For locally advanced disease, chemoradiotherapy (CRT) improves survival compared with radiotherapy (RT) alone in selected populations, yet this benefit is heterogeneous and depends on tumor burden, disease stage, and patient fitness. Population-average estimates from trials and observational studies provide essential guidance but do not capture the substantial inter-patient variability observed in clinical practice (1,2).

Digital-twin frameworks have been proposed as a means to simulate patient-specific outcomes under alternative treatment strategies (3,4). However, many existing approaches in oncology focus on predictive modeling of risk or prognosis without an explicit causal framework, which limits their ability to support counterfactual reasoning about how outcomes would change under alternative treatment decisions. Causal inference methods, including doubly robust learners and causal forests, provide a principled framework for estimating individualized treatment effects from observational data by accounting for confounding and treatment-selection bias (5,6). Their integration with interpretable feature-selection strategies and policy-level evaluation remains limited in radiation oncology (7).

In this work, we present a causal digital-twin framework for H&N cancer that combines Bayesian-network (BN)–derived Markov Blankets (MB) with doubly robust individualized treatment-effect estimation to simulate patient-specific survival benefit from CRT versus RT. Analyses are performed separately for human papillomavirus (HPV)-negative and HPV-positive disease using a temporally separated training and test design. We evaluate treatment-effect heterogeneity and compare τ□-guided selective treatment policies against uniform strategies, emphasizing counterfactual simulation and decision relevance rather than risk prediction alone.

## Materials and Methods

### Data and cohort

We analyzed the public RADCURE (8) clinical cohort (n=3,346) and stratified patients by HPV status into HPV-negative and HPV-positive subcohorts. For each subcohort, patients were split temporally into training (RT start year ≤2007) and held-out test sets (RT start year >2007) to assess out-of-time generalization.

### Endpoint

The primary endpoint was binarized 2-year survival (SVy2), computed from RT start to date of death (if available) or last follow-up, and labeled SVy2=1 for survival ≥24 months SVy2=0 otherwise.

### Causal digital-twin framework

For each HPV-stratified cohort, a BN structure-learning workflow was used to identify the MB of SVy2, yielding a minimal, interpretable covariate set for downstream modeling (full preprocessing/MB details described in our prior work (9)). Using MB covariates, we estimated individualized treatment effects of CRT vs RT alone with a doubly robust causal forest (DML), targeting. Propensity scores were fit on training data and used to enforce overlap (common support) to reduce extrapolation. We report test-set ATE (mean), ITE summaries, and policy evaluation using doubly robust (AIPW) estimates of policy value for selective-CRT strategies compared with treat-all and treat-none baselines. Figure 1 summarizes the overall framework and evaluation pipeline.

**Figure 1.**
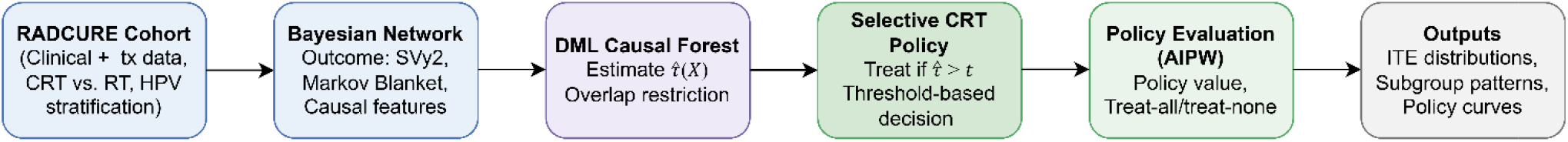
Overview of the proposed causal digital-twin framework. Bayesian network–based causal feature selection is followed by doubly robust causal forest estimation of individualized treatment effects, enabling selective chemoradiotherapy policies and policy-level evaluation.

## Results

### Prognostic performance using Markov Blanket features

In the HPV-negative cohort, the Markov Blanket–based prognostic model demonstrated good discrimination on the temporally held-out test set (AUC = 0.72; C-index = 0.81, log-rank p < 0.01; Fig. 2a and 2b). Kaplan–Meier analysis using a training-derived risk threshold showed clear separation between low- and high-risk groups over follow-up. In contrast, prognostic performance in the HPV-positive cohort was attenuated (test AUC = 0.61; C-index = 0.65), with no statistically significant survival separation (log-rank p = 0.11; Fig. 2c and 2d), consistent with the more uniformly favorable prognosis of HPV-positive disease

**Figure 2.**
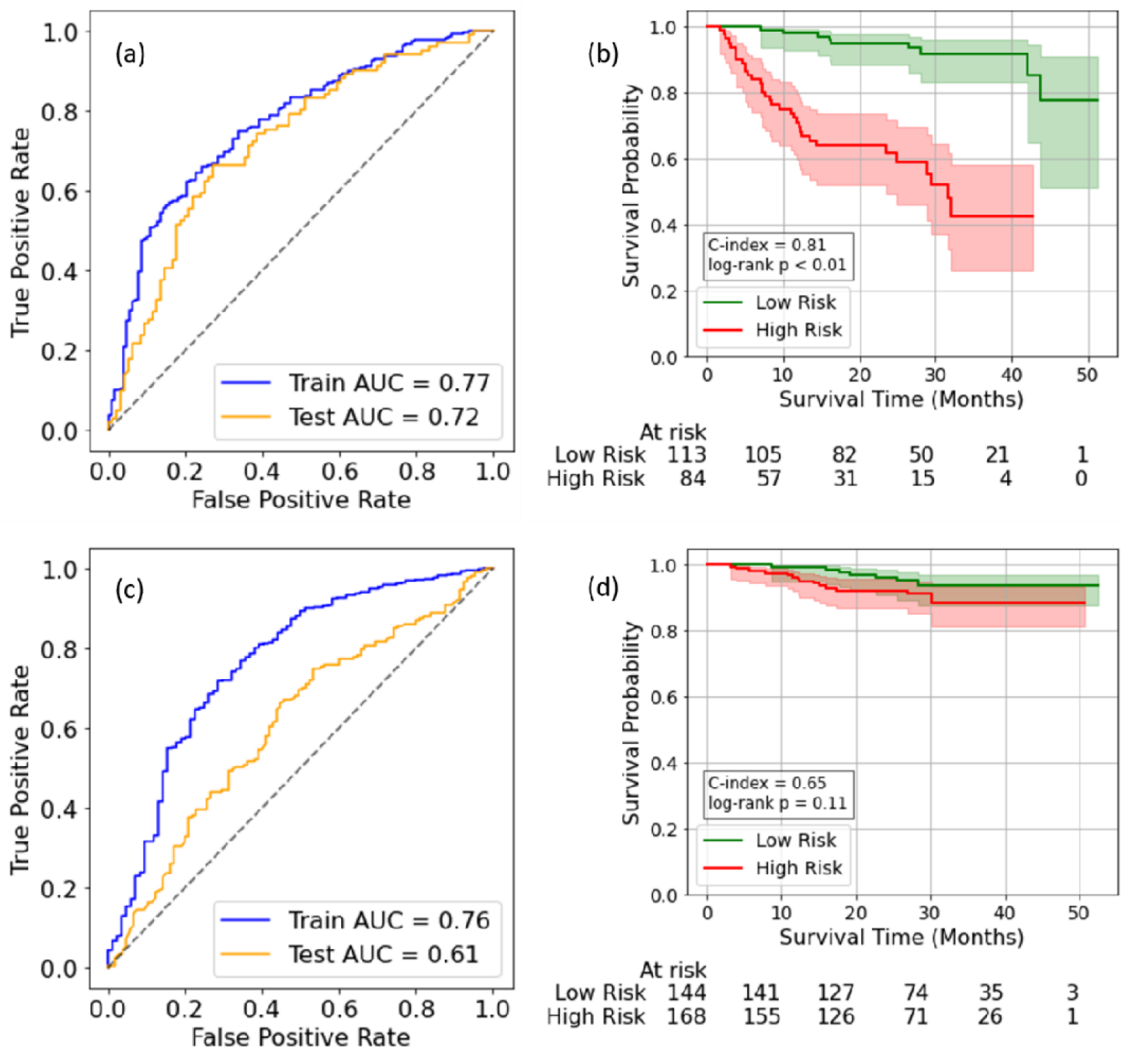
Prognostic discrimination of Markov Blanket–based models by HPV status. **(a–b)** HPV-negative cohort shows good test-set discrimination (AUC = 0.72) and significant Kaplan–Meier risk separation (C-index = 0.81, log-rank p < 0.01). **(c–d)** HPV-positive cohort shows reduced discrimination (AUC = 0.61) and attenuated, non-significant survival separation (C-index = 0.65, log-rank p = 0.11). Risk thresholds were derived from training data and applied unchanged to the test set.

### Individualized treatment-effect estimation

In HPV-negative patients, causal forest estimation yielded a modest but positive average treatment effect favoring CRT over RT (ATE = 0.136; 95% CI [0.124, 0.148]). The distribution of individualized treatment effects (ITE,) was right-shifted with limited variance (SD = 0.086), and most patients exhibited small-to-moderate estimated benefit (q10 = 0.006, q90 = 0.251; Fig. 3a). Clinically coherent patterns were observed, with higher among patients with advanced nodal disease and good performance status, and lower benefit in patients with poorer ECOG status (Fig. 4). In HPV-positive patients, the mean estimated treatment effect was similar in magnitude (ATE = 0.139; 95% CI [0.035, 0.237]) but was accompanied by substantially greater heterogeneity (SD = 0.93), with both large positive and negative values (Fig. 3b). No consistent stratification of by nodal stage or ECOG performance status was observed, suggesting limited clinical structure in treatment-effect heterogeneity within this subgroup.

**Figure 3.**
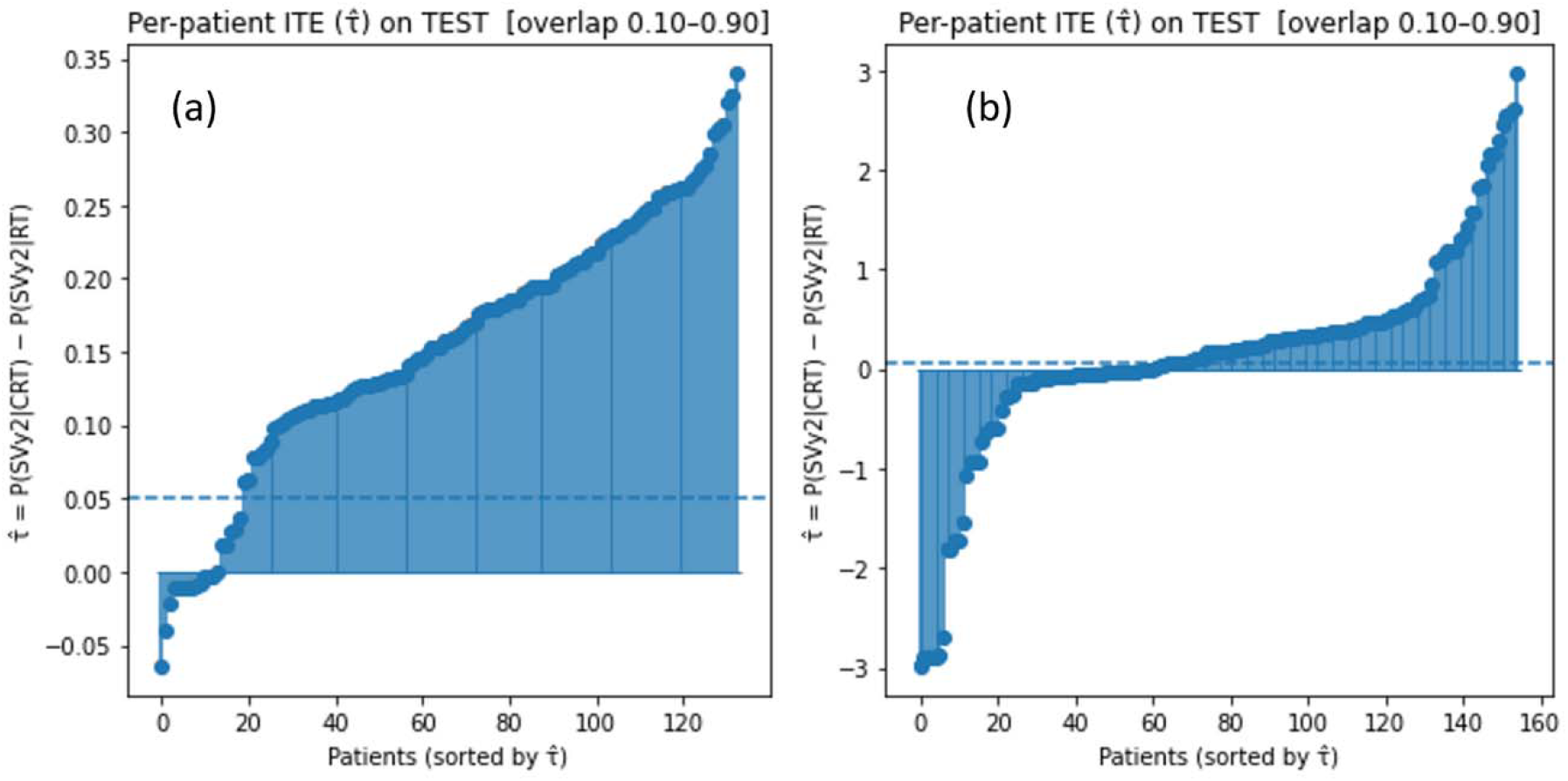
Per-patient ITEs () for CRT versus RT in the held-out test set, restricted to the region of adequate propensity overlap (0.10–0.90). Patients are sorted by within each cohort: **(a)** HPV-negative and **(b)** HPV-positive disease. The dashed line denotes a representative clinical decision threshold ().

**Figure 4.**
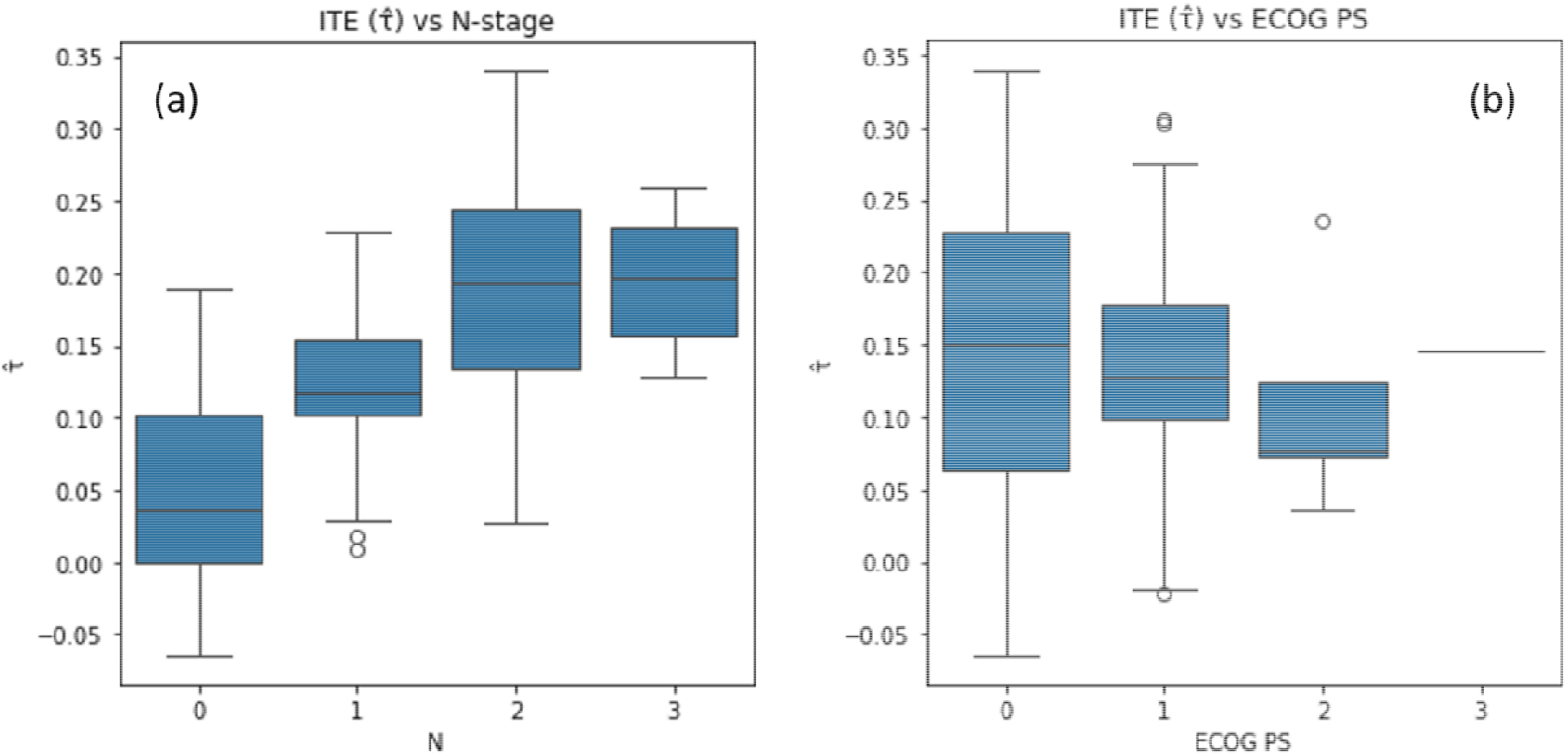
ITEs () for CRT versus RT in the HPV-negative cohort stratified by **(a)** nodal stage (N) and **(b)** ECOG performance status (PS). Boxes denote interquartile ranges with medians; whiskers indicate range.

### Policy evaluation and decision analysis

For the HPV-negative cohort, a selective CRT policy based on thresholds yielded modest improvements in estimated SVy2 compared with uniform treatment strategies (policy value 0.429 vs 0.426 for treat-all and 0.439 for treat-none; Fig. 5a). Using a clinically meaningful threshold (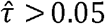), the model recommended CRT for 80.2% of test patients.

**Figure 5.**
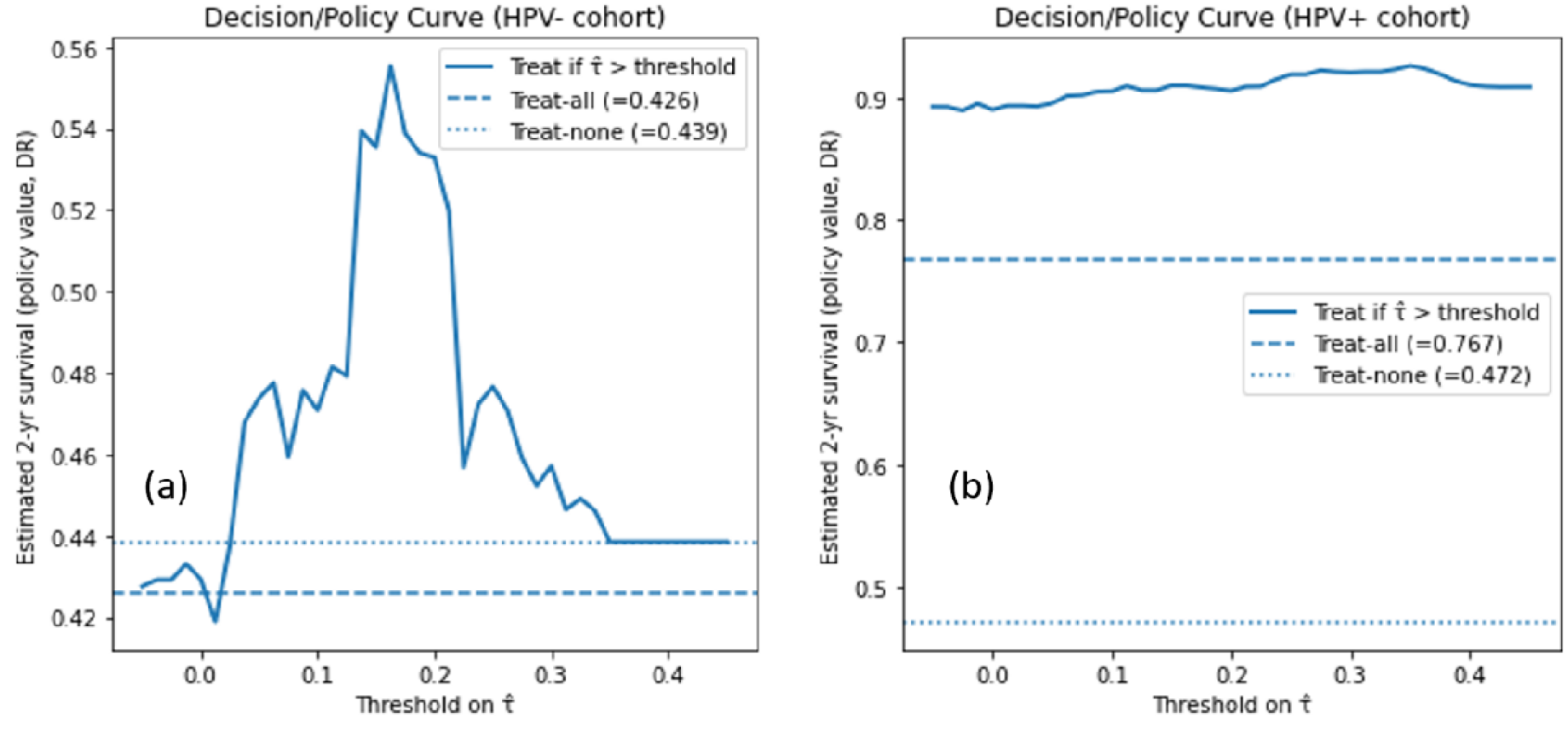
Decision (policy) curves comparing selective CRT policies based on individualized treatment effects () with treat-all and treat-none strategies in **(a)** HPV-negative and **(b)** HPV-positive cohorts. Policy value represents estimated SVy2 under each decision rule using a doubly robust estimator.

In the HPV-positive cohort, selective 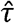-guided treatment policies achieved substantially higher estimated policy value than uniform strategies (policy value 0.890 vs 0.767 for treat-all and 0.472 for treat-none; Fig. 5b). However, these gains were driven by a small subset of patients with large, estimated effects and occurred in the setting of wider uncertainty and reduced overlap, and should therefore be interpreted as exploratory.

## Discussion

In this study, we present a causal digital-twin framework for estimating individualized survival benefit from CRT versus RT in H&N cancer, stratified by HPV status. By integrating BN–based MB selection with doubly robust causal forest estimation, the framework moves beyond risk prediction toward counterfactual, policy-relevant treatment simulation. For HPV-negative disease, the model identified clinically coherent heterogeneity in CRT benefit. ITE () were generally positive but varied substantially across patients, with larger estimated benefit in node-positive disease and in patients with better performance status. Importantly, policy evaluation demonstrated that selective CRT guided by could achieve comparable or improved estimated SVy2 relative to uniform treatment strategies, despite modest differences between treat-all and treat-none baselines. These findings suggest that individualized treatment-effect modeling may help identify subgroups in whom treatment intensification is most likely to confer meaningful benefit.

In contrast, HPV-positive disease exhibited markedly different behavior. Although the average treatment effect favored CRT, the distribution of was substantially wider, with extreme positive and negative estimates and reduced overlap between treatment groups. Policy curves showed higher estimated survival for -guided strategies compared with uniform policies; however, this apparent gain should be interpreted cautiously given limited overlap, wide uncertainty, and the uniformly favorable prognosis of HPV-positive disease. Together, these results are consistent with the hypothesis that treatment-effect heterogeneity in HPV-positive patients is more difficult to estimate reliably from observational data and may not translate into stable, actionable decision rules.

Several strengths distinguish this work. First, causal feature selection via MBs yielded compact, interpretable models aligned with known clinical factors. Second, temporal train–test separation reduced information leakage and better approximated prospective deployment. Third, evaluation emphasized policy-level metrics rather than predictive accuracy alone, aligning model assessment with clinical decision-making.

This study also has limitations. As an observational analysis, causal estimates rely on standard assumptions of no unmeasured confounding and sufficient overlap, which were imperfect— particularly in the HPV-positive cohort. Treatment intensity, toxicity, and quality-of-life outcomes were not modeled, and external validation was not performed in this submission. These limitations motivate future work incorporating multi-institutional validation, toxicity-aware utility functions, and explicit uncertainty quantification.

In conclusion, this work demonstrates the feasibility of a causal digital-twin framework for individualized treatment-effect estimation in radiation oncology. The results highlight clinically meaningful heterogeneity in CRT benefit for HPV-negative disease while underscoring the challenges of individualized policy learning in HPV-positive patients. With external validation and extension to multi-objective decision support, such frameworks may provide a principled foundation for personalized treatment selection beyond population-average recommendations.

## Data Availability

All data produced in the present study are available upon reasonable request to the authors

https://www.cancerimagingarchive.net/collection/radcure/

## Author Contributions

KDS: Conceptualization, Data curation, Formal analysis, Methodology, Software, Visualization, Validation, Writing-original draft; CWC, JZ, JD, AD, HAA, JB, WS, DSY: Writing-review & editing; XY: Funding acquisition, Supervision, Validation, Writing-review & editing

## Acknowledgments

We would like to acknowledge The Cancer Imaging Archive (TCIA) for hosting the RADCURE data set and the University Health Network (UHN), Toronto, Canada, for providing and curating the data used in this study.

